# Identifying Cardiogenic Shock Sub-Phenotypes with Machine Learning: A Multicenter Study Combining Clinical and Echocardiographic Data

**DOI:** 10.1101/2025.04.28.25326615

**Authors:** Nicolò Ghionzoli, Andrea Stefanini, Geza Halasz, Carlotta Sorini Dini, Carlotta Sciaccaluga, Francesca Maria Righini, Valerio Guarrasi, Arianna Francesconi, Paolo Soda, Maria Concetta Pastore, Hatem Soliman Aboumarie, Vito Piazza, Marco Marini, Domenico Gabrielli, Matteo Cameli, Serafina Valente

## Abstract

**Background:** Sub-phenotyping cardiogenic shock (CS) patients using non-traditional clustering methods represents a step toward precision medicine, potentially improving outcomes in this heterogeneous and high-mortality condition. This study aimed to apply an unsupervised machine learning approach to integrate clinical and advanced echocardiographic data, identifying CS sub-phenotypes associated with different outcomes and features, beyond etiology.

**Methods:** This multicenter observational study prospectively analyzed 172 patients admitted to Cardiac Intensive Care Units with overt CS, from 2021. An exploratory statistical analysis preceded patient clustering using the Elbow Method and K-Means algorithm, based on clinical presentation. Dimensionality reduction was performed with Principal Component Analysis. Phenotypes were further stratified according to the Society for Cardiovascular Angiography and Interventions (SCAI) stages.

**Results:** Five distinct Phenotypes (labeled from I to V) were identified, showing progressively increasing in-hospital mortality rates: 25% (I), 32% (II), 39% (III), 41% (IV), and 60% (V). Kaplan-Meier analysis demonstrated a stepwise increase in mortality risk. Phenotypes IV and V had significantly higher mortality than Phenotype I (HR: 2.78 [95% CI, 1.07-7.19] and HR: 2.80 [95% CI, 1.10-7.14]; P < 0.05). Mortality prediction remained independent after adjustment for confounding factors, and independently of SCAI stage. Phenotype I had the lowest mortality, with higher arterial pressure and moderate left ventricular (LV) dysfunction, whereas Phenotype II exhibited marked LV failure. Oppositely, Phenotypes IV and V had severe congestion despite only mild LV impairment.

**Conclusions:** Machine learning, newly integrating echocardiographic data, identified five distinct CS Phenotypes, each with unique clinical/echocardiographic features and mortality risks. These insights could support personalized treatment strategies in CS patients, pending further validation.

**WHAT IS NEW?:** This machine learning-based analysis, newly integrating echocardiographic data, identified 5 distinct CS Phenotypes. These Phenotypes have been shown to be easily feasible at the bedside, each with unique mortality risks and features, including varying patterns of cardiac dysfunction and dilatation.

**WHAT ARE THE CLINICAL IMPLICATIONS?:** The 5 CS Phenotypes lay the foundation for personalized treatment strategies and improve risk stratification beyond SCAI staging in CS patients. Further external validation of the 5 Phenotypes to other independent CS cohorts, and additional investigations that account for trajectories of patients across Phenotypes, could establish their role in enhancing clinical outcomes.

## INTRODUCTION

Cardiogenic shock (CS) is a subtype of circulatory shock in which end-organ perfusion has inadequate cardiac output as *primum movens*. It is characterized by the need for vasoactive support and clinical signs of tissue hypoperfusion, together with increased serum lactate levels, resulting in a wide spectrum of phenotypes that reflect diverse etiologies, pathogenetic mechanisms, stages of severity and patient-specific conditions [1–4].

Despite advancements in management, including mechanical circulatory supports, CS remains associated with an extremely high in-hospital mortality rate, ranging from 30% to 60% [1, 2, 5, 6]. As the public health burden of CS is expected to rise over time, several prognostic scores and classifications have been formulated, in order to best allocate available resources and appropriate therapeutic strategies [7–12]. Specifically, the Society for Cardiovascular Angiography and Interventions (SCAI) proposed a five-stage classification of CS, validated in retrospective studies and recently updated, based on clinical assessment of hypoperfusion, lactate levels, and hemodynamic evaluation [12–14]. Additionally, the sub-phenotyping of patients with CS continues to evolve to disentangle heterogeneity, and the use of an impartial machine learning (ML)-based clustering algorithm – which simplifies routine clinical data into reproducible CS sub-phenotypes – may yield new insights that could support more personalized care [15]. This approach has been effectively applied for the phenotyping of several clinical syndromes and disorders [16, 17], including CS, with the identification of distinct phenotypes, each with specific and reproducible correlations to mortality [18, 19].

All these classifications include both hemodynamic variables and indices of metabolic derangement, but lack echocardiographic data, which is the promptest and safest diagnostic modality [2, 20], with relevant prognostic role [21–26]. An integrative approach combining clinical variables and echocardiographic parameters could help identify differences in underlying disease mechanisms, offering insights with prognostic and therapeutic implications.

We primary aimed to apply an unsupervised ML approach integrating clinical and imaging data (including advanced echocardiography), to identify clinically relevant CS sub-phenotypes associated with different outcomes and treatment responsiveness, as a promising step forward in precision medicine, beyond etiology. As for the secondary objectives, we aimed to characterize the clinical profiles and outcomes associated with identified sub-phenotypes and their reproducibility. Finally, we sought to improve risk stratification of CS patients, in particular by defining subsets of mortality risk within the SCAI staging system.

## METHODS

### STUDY DESIGN AND POPULATION

In this prospective, observational, multicenter, non-profit study, patients diagnosed with CS at admission to the three Italian cardiac intensive care units were enrolled from November 1st, 2021 to September 30th, 2024. CS diagnosis was physician adjudicated at each site based on the European Society of Cardiology guidelines [3]. In particular CS was diagnosed based on the presence of clinical signs of hypoperfusion (altered mental status, oliguria, cold extremities, narrow pulse pressure, dizziness) associated with biochemical markers of hypoperfusion (elevated creatinine, metabolic acidosis and elevated serum lactate), with or without hypotension (defined as systolic blood pressure < 90 mmHg for more than 30 min, or the need for catecholamines to maintain systolic blood pressure > 90 mmHg) [3].

Thus, patients underwent clinical, biohumoral, and echocardiographic evaluation at admission, within 3 hours from CS identification. Then, all the following clinical events were registered during the course of patients’ hospital stay: in-hospital mortality, major arrhythmias, use of temporary mechanical circulatory supports, heart transplantation and/or left ventricular (LV) assist devices implantation.

Only adult patients (aged ≥ 18 years) were considered for analysis, and patients with incomplete data for key variables (in particular missing outcome data) were excluded. All patients provided written informed consent and the study was conducted in accordance with the Declaration of Helsinki. The study was approved by the Tuscany Region Local Ethic Committee for Clinical Trial – section south-west extended area (*approval number: 20438*). The corresponding author had full access to all the study data and takes responsibility for its integrity and the data analysis.

### CLINICAL, LABORATORISTIC AND ECHOCARDIOGRAPHIC PARAMETERS

For each patient the following data were collected and extracted from electronic health records: baseline demographic characteristic, cardiovascular risk factors, number of vessels affected and culprit lesion (if applicable) if acute coronary syndrome (ACS) etiology, vital signs, cardiac arrest as onset, principal drugs used during hospitalization, blood tests at admission (including blood count, C-reactive protein, kidney function, liver function, high-sensitivity troponin, N-terminal pro B-type natriuretic peptide, arterial lactate levels, arterial oxygen saturation, central venous oxygen saturation), and the use of mechanical circulatory supports.

Echocardiography was performed by an expert operator using a fully equipped machine (Vivid E9, GE, Horthern, Norway). All parameters were measured according to the European Association of Cardiovascular Imaging/American Society of Echocardiography guidelines [27] [*Supplemental methods*]. Speckle tracking echocardiography was performed offline using the dedicated 2D strain softwares (Echopac, GE), by a single independent operator, for each center who analyzed all the images acquired by a second experienced operator in the same center [*Supplemental methods*].

### VARIABLE HANDLING AND CLUSTER ANALYSIS

An initial statistical analysis focused on candidate variables and in-hospital mortality correlations, using Pearson’s correlation coefficient, was applied. Results were visualized through heatmaps, highlighting the strength and direction of these relationships. Hence, a careful selection of included variables for the subsequent clustering was based not only on their associations with in-hospital mortality, but also on clinical judgement, reflecting underlying disease and pathophysiological processes, in order to avoid the identification of clusters based on outcomes rather than pathophysiology. A sensitivity analysis was conducted by testing clustering stability with and without specific variables (LV global longitudinal strain (GLS), lactate, right ventricular fractional area change). Patients with high proportions of missing data were excluded, to ensure that overall missingness did not exceed 20% for imputation. Remaining missing values were imputed using the K-Nearest Neighbors Imputer (K = 5). The number of variables was established according to the simple size (considering the Formann formula in which clustering analysis should include 2^*n*^ individuals, where n is the number of variables) [15].

Following the exploratory analysis, clustering was conducted by an external expert using the K-Means algorithm. The number of clusters was optimized using the Elbow Method by evaluating the sum of squared distances and further validated with the Silhouette Score, which measures clustering quality. Each cluster’s distinctiveness was examined in relation to in-hospital mortality as the outcome variable using Chi-square test. Dimensionality reduction was performed using Principal Component Analysis to facilitate the visualization of clusters in a two-dimensional space. The Principal Component Analysis plot highlighted the distribution of clusters and their alignment with the target variable.

### CONVENTIONAL/TRADITIONAL STATISTICAL ANALYSIS

All continuous variables were tested for normality using the Kolmogorov-Smirnov test. Continuous variables were expressed as mean ± standard deviation or median and interquartile ranges (IQR), and depending if two-group or five-group comparison, data were compared using the Student’s t-test/Analysis of Variance (ANOVA) or Mann-Whitney/Kruskal-Wallis test, as appropriate. Categorical data were expressed as counts and percentages and were compared using the Chi-square test. All features within each Phenotype were described and compared to summarize each cluster’s key characteristics and all phenotypes were further stratified according to the SCAI shock stage at admission.

Univariate and multivariate Cox regression analysis was also used to assess independent predictors of in-hospital mortality, with adjustment for confounding factors (age, sex, CS etiology and mechanical circulatory supports). To examine the impact of phenotype membership on in-hospital mortality, survival curves were generated by the Kaplan–Meier estimator and compared using log-rank test. Analyses were performed by an external expert using the Statistical Package for Social Sciences software, release 30.0 (SPSS). A P value < 0.05 was considered statistically significant.

## RESULTS

### STUDY POPULATION

Overall, 172 CS patients were enrolled (mean age 65 ± 14 years, 78% (135 patients) male, and BMI 26.4 ± 4.5 kg/m^2^). Nearly half of patients had a history of hypertension (54%) and dyslipidemia (56%) and the use of at least one mechanical circulatory support was frequent: 52% (89 patients) with intra-aortic balloon pump, 15% (26) with Impella and 14% (23) with extracorporeal membrane oxygenation. Overall in-hospital mortality was 34%. The most common cause of CS was ACS (56%, 97 patients), with non-ACS etiologies accounting for the remaining 44% (75 patients). As for echocardiographic parameters, the population showed enlarged left ventricle (LV end-diastolic diameter (EDD): 57 ± 11 mm) and left atrium (left atrial volume: 76 ± 17 ml), and severe LV dysfunction as measured by LV ejection fraction (EF) of 22 (15-30) %. Right ventricular global function was nearly normal (S’ tissue Doppler imaging (TDI): 0.09 (0.07-0.12) m/s, right ventricular fractional area change: 32 (25-40) %). Regarding STE parameters, patients showed a reduction in LV strain, with LV GLS -6.1 ± 3.5%. The general characteristics of the overall study population are reported in *Table 1*. No sex-based differences were present. The overall missingness of the data was 18%; LV GLS was the feature with the highest missingness (50%), reflecting daily practice challenges in cardiac intensive care unit imaging.

**Table 1.**
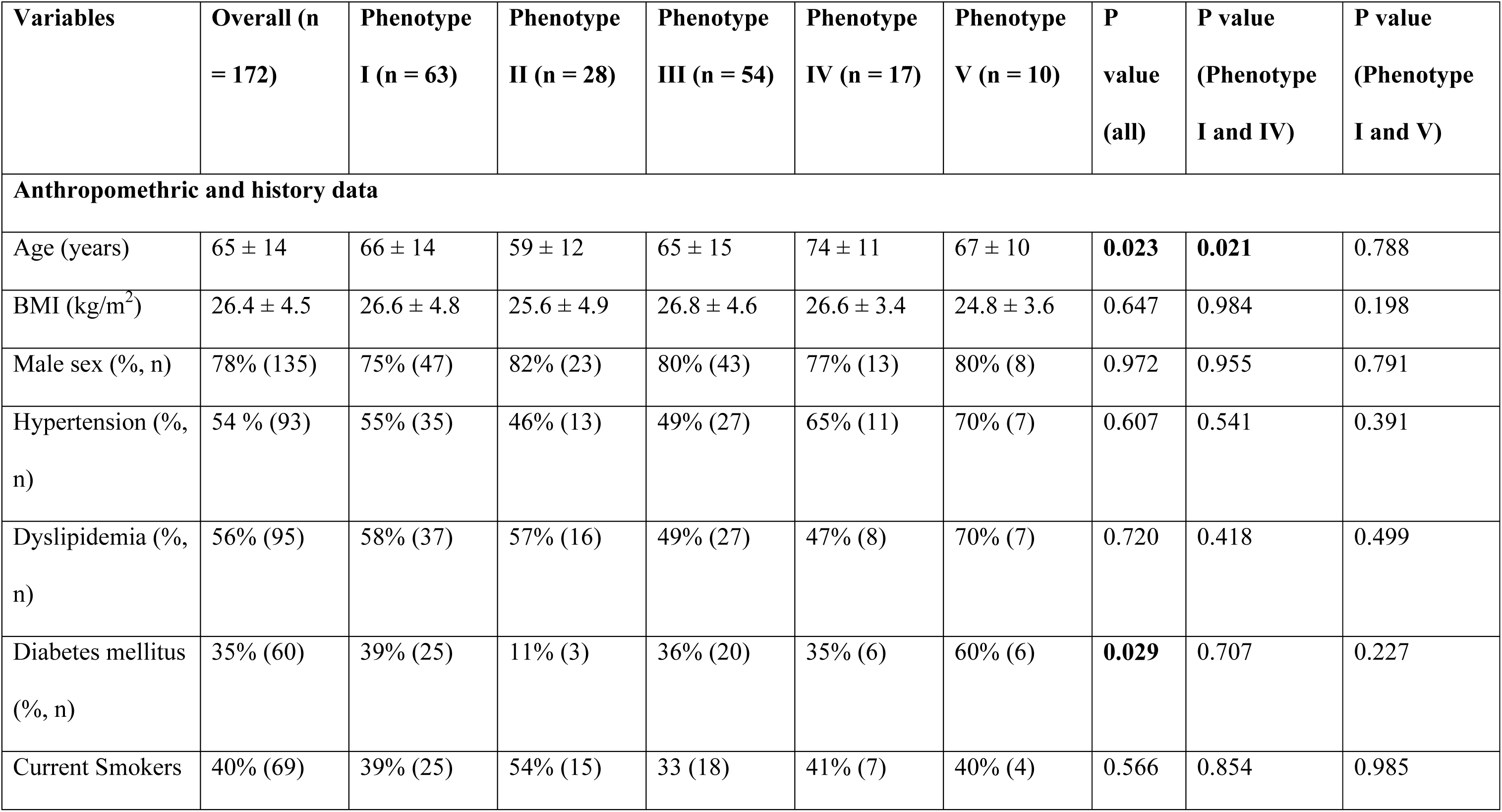
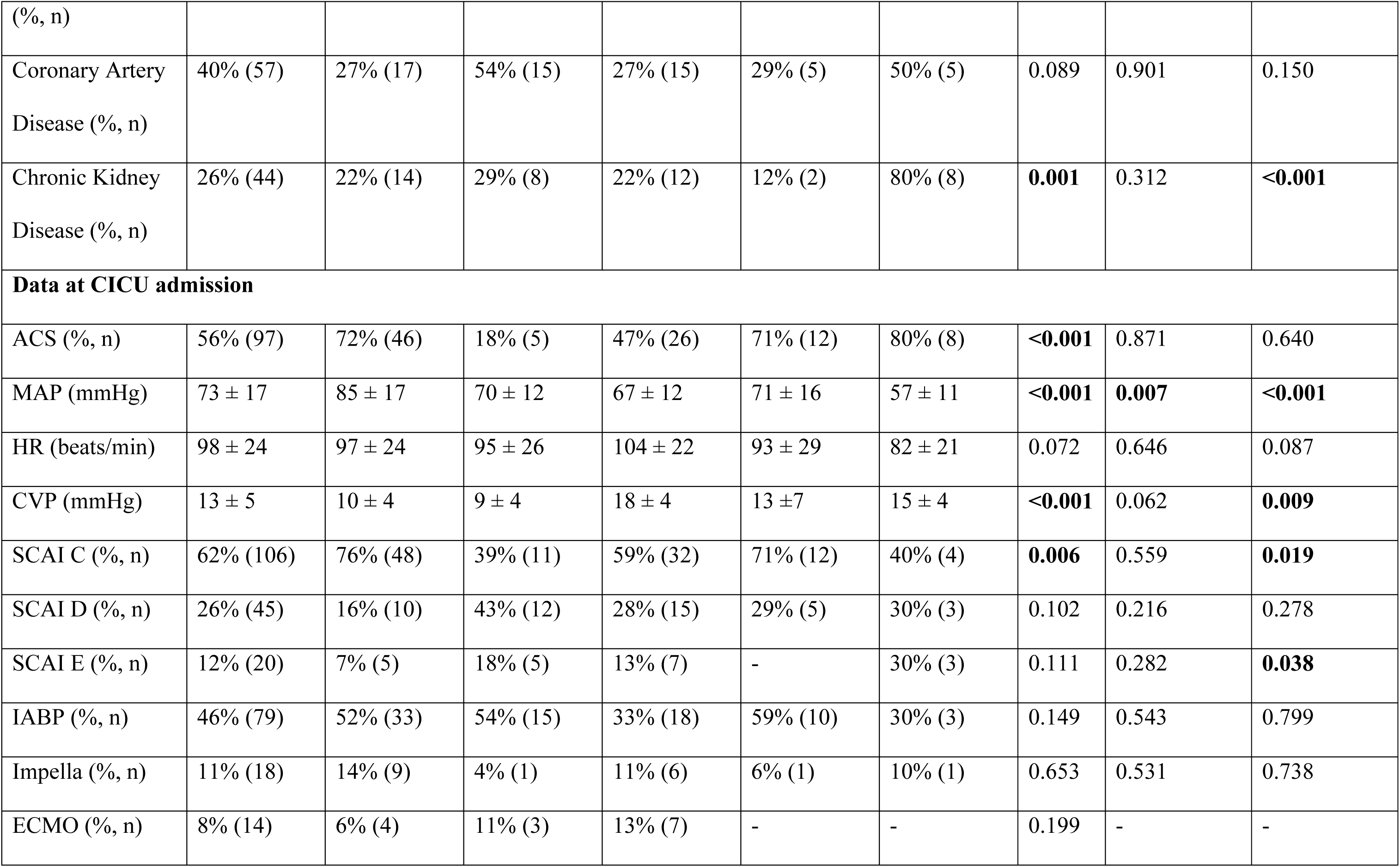
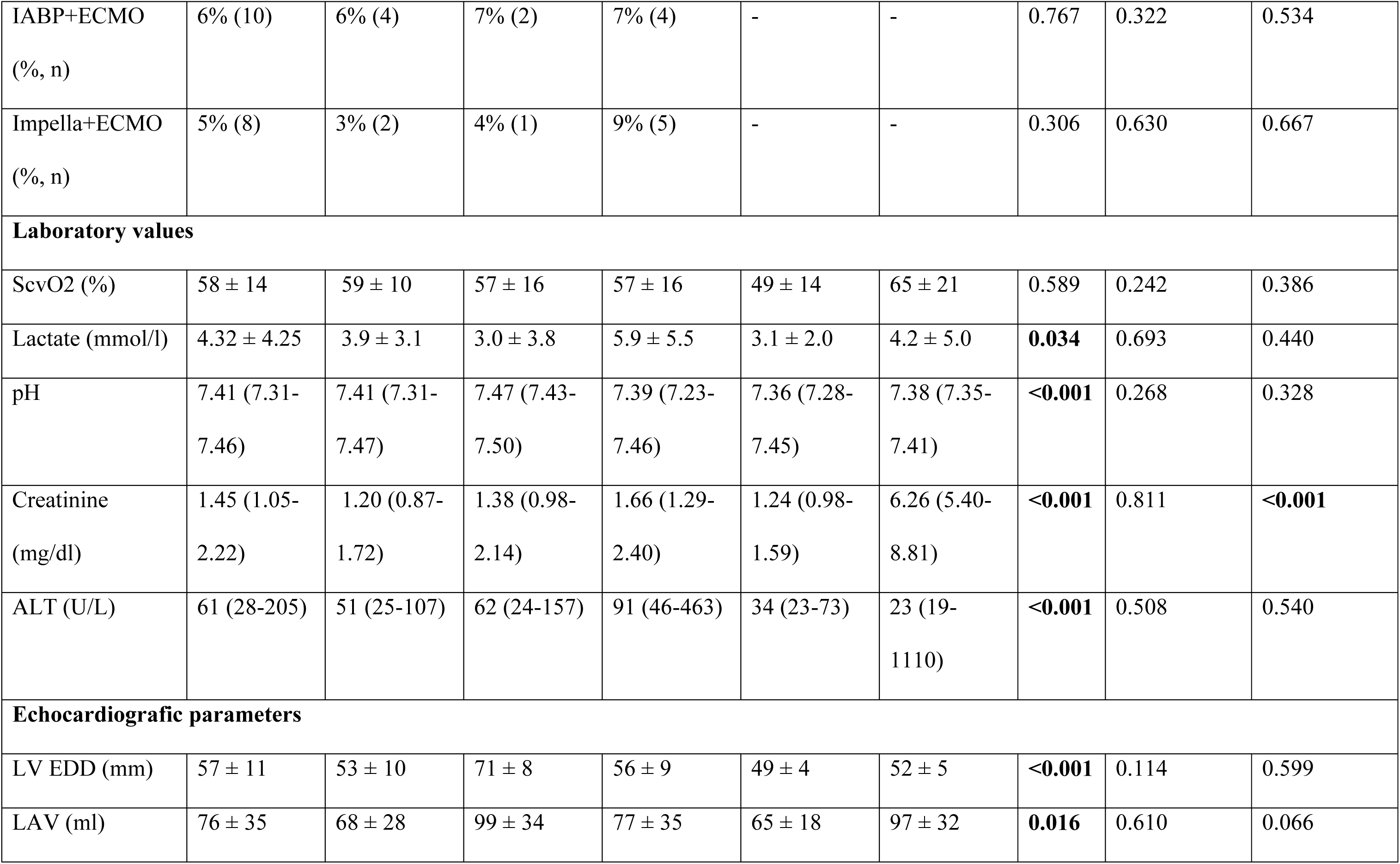
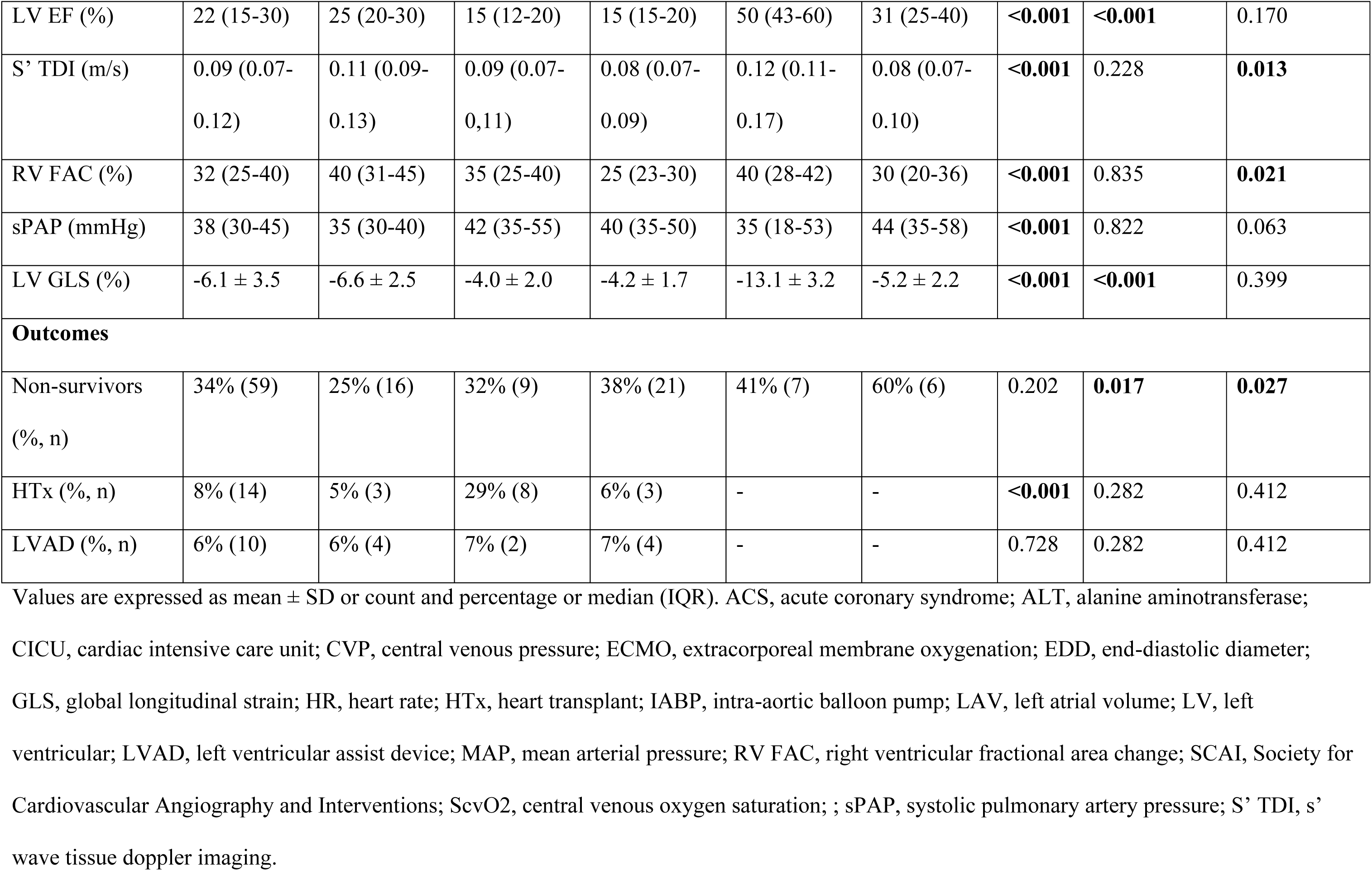
Characteristics and outcomes of the overall study population and by Phenotypes at the time of admission or during the Hospital stay.

### PHENOTYPES OF CARDIOGENIC SHOCK

After calculating Pearson’s correlation coefficient, age, creatinine, lactate level, procalcitonin and C-reactive protein showed the greatest association with in-hospital mortality (*Figure S1*). Then, following a careful selection to avoid the identification of clusters based on outcomes rather than pathophysiology, 8 final variables were included in the clustering analysis, including echocardiographic parameters: mean arterial pressure (MAP), central venous pressure, lactate, creatinine, LV EDD, LV EF, S’ TDI and LV GLS.

An optimal cluster number of 5 was determined (*Figure S2*).

K-means algorithm identified 5 CS Phenotypes labeled from I to V (*Figure 1*) based on their unique clinical and echocardiographic characteristics at presentation (*Figure 2*, *Table 2*). The Principal Component Analysis plot highlighted the distribution of the 5 clusters and their alignment with the target variable (*Figure S3*). Radar plot was used to display the deviation of clinical and echocardiographic values from the mean value (*Figure 3*). Baseline key characteristics and outcomes across Phenotypes are summarized in *Table 1*. Phenotype I, of intermediate age (66 ± 14 years) and lowest in-hospital mortality, had a preserved MAP (85 ± 17 mmHg), nearly normal kidney function (creatinine: 1.20 (IQR 0.87-1.72) mg/dl) and a slight increase in LV EDD (53 ± 10 mm, P = 0.006) with a moderate reduction in LV EF and LV GLS (respectively 25 (IQR: 20-30) % and -6.6 ± 2.5%; P < 0.001) (with LV GLS intraclass correlation coefficient: 0.99 (IQR: 0.99-0.99). Phenotype II exhibited similar metabolic and hemodynamic features relative to Phenotype I, but, interestingly, marked LV dilation (EDD: 72 (IQR: 66-76) mm; P < 0.001) and dysfunction (LV EF: 15 (IQR: 12-20) %; P < 0.001) and preserved right ventricular function (S’ TDI: 0.09 (IQR 0.07-0,11) m/s). Phenotype III, of intermediate age (65 ± 15 years) and in-hospital mortality, primarily showed RV dysfunction (right ventricular fractional area change: 25 (IQR 23-30) %, P < 0.001; S’ TDI: 0.08 (IQR: 0.07-0.09) m/s, P < 0.001; free wall right ventricular longitudinal strain: -13.5 ± 5.9%, P = 0.003) and a higher grade of lactate level (5.9 ± 5.5 mmol/l), systemic congestion (central venous pressure: 18 ± 4 mmHg, P < 0.001) and renal dysfunction (creatinine: 1.66 (1.29-2.40) mg/dl). Conversely, Phenotype IV, of older age (74 ± 11 years, P = 0.003), and Phenotype V showed the highest in-hospital mortality associated with elevated lactate, relevant systemic congestion (central venous pressure: 13.0 ± 7 mmHg and 15.0 ± 4 mmHg, respectively) but only a mild reduction in LV EF (50 (IQR 43-60) % and 31 (IQR 25-40) %, for each), with preserved LV dimension (EDD: 49 ± 4 mm and 52 ± 5 mm, respectively) and right ventricular function. In addition, Phenotype V exhibited severe hypotension (MAP: 57 ± 11 mmHg; P < 0.001) associated with severe renal dysfunction (creatinine: 6.26 (IQR 5.40-8.81) mg/dl; P < 0.001).

**Figure 1.**
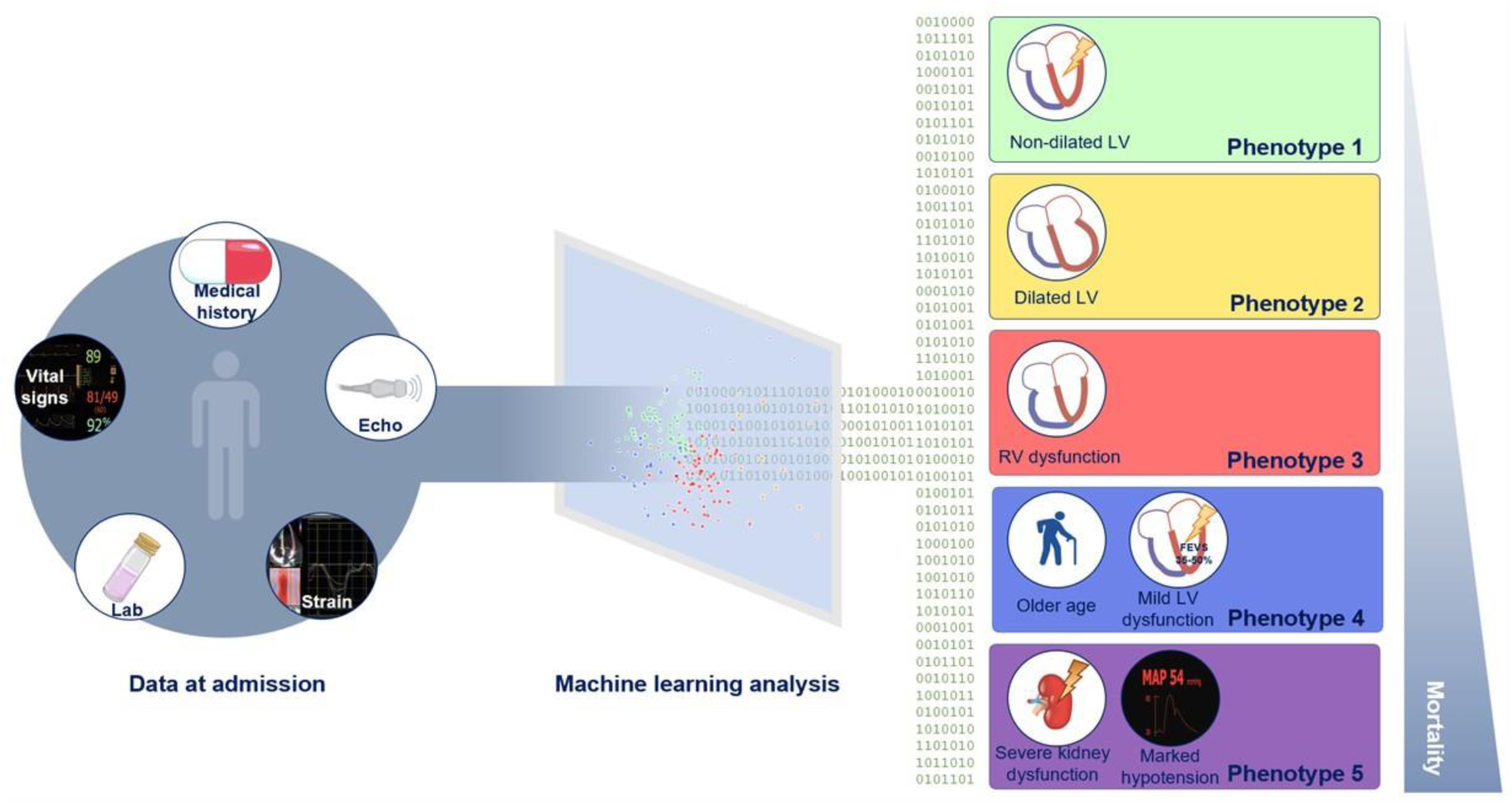
Machine learning and Echo Data for Cardiogenic Shock Phenotypes - Machine Learning-based integration of clinical and echocardiographic multicenter data to identify 5 cardiogenic shock Phenotypes, each with unique mortality risks and features. LV, left ventricular; MAP, mean arterial pressure; RV, right ventricular.

**Figure 2.**
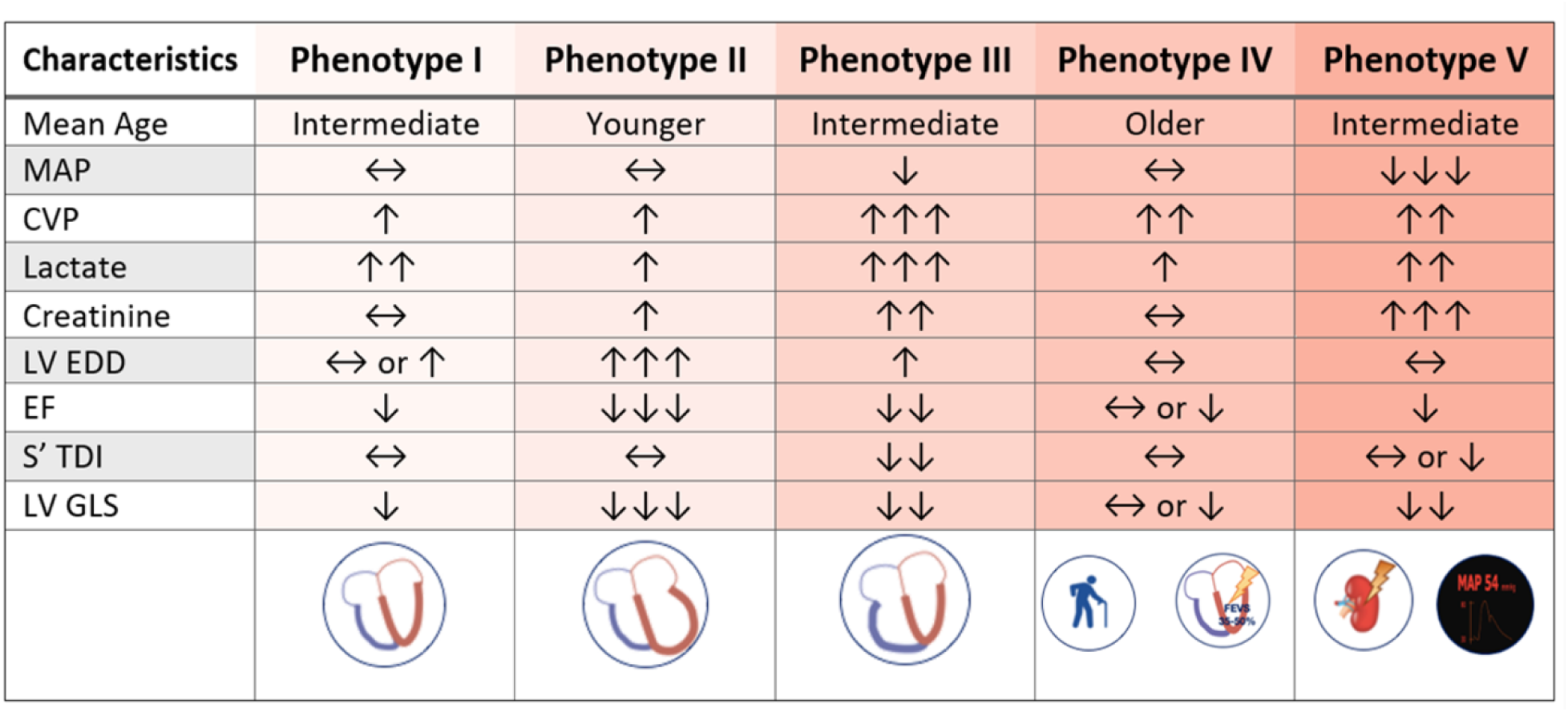
The 5 distinct cardiogenic shock Phenotypes - Outstanding characteristics at presentation of the 5 cardiogenic shock Phenotypes. CVP, central venous pressure; EDD, end-diastolic diameter; EF, ejection fraction; GLS, global longitudinal strain; LAC, lactate; LV, left ventricular; MAP, mean arterial pressure; S’ TDI, s’ wave tissue doppler imaging.

**Figure 3.**
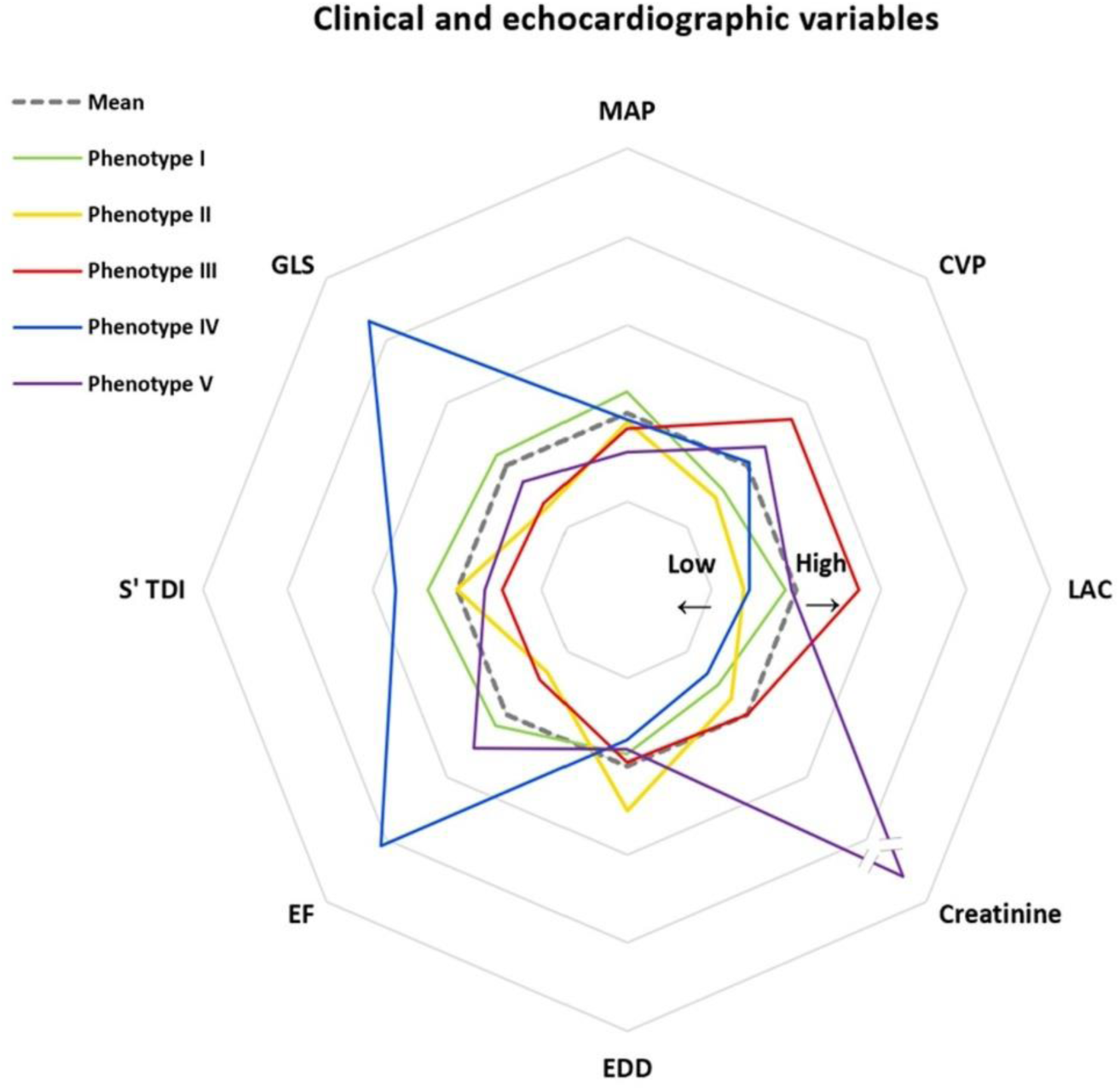
Phenotypes’ clinical and echocardiographic trends **–** This radar plot illustrates the distribution for each Phenotype of clinical and echocardiographic variables. Data were normalized across all Phenotypes. The dashed gray line marks the mean values. Values above the mean are drawn outside the dashed line, whereas values below are drawn inside. The creatinine value scale for Phenotype V has been reduced (as indicated by the double lines on the purple line) to better adjust the relationships between the other Phenotypes and the mean. CVP, central venous pressure; EDD, end-diastolic diameter; EF, ejection fraction; GLS, global longitudinal strain; LAC, lactate; MAP, mean arterial pressure; S’ TDI, s’ wave tissue doppler imaging.

**Table 2.** Peculiar and distinguishing features of each Phenotype compared to the others.

| **Variable** | **Phenotype** | **Other phenotypes** | **P value** |
| --- | --- | --- | --- |
|  | **Phenotype I** | **Non-phenotype I** |  |
| LV EDD (mm) | 53 ± 10 | 60 ± 11 | **0.001** |
| LV EF (%) | 25 (20-30) | 20 (15-25) | **<0.001** |
|  | **Phenotype II** | **Non-phenotype II** |  |
| LV EDD (mm) | 71 ± 8 | 54 ± 9 | **<0.001** |
|  | **Phenotype III** | **Non-phenotype III** |  |
| S' TDI (m/s) | 0.08 (0.07;0.09) | 0.11 (0.08-0.12) | **<0.001** |
| RV FAC (%) | 25 (23-30) | 37 (30-42) | **<0.001** |
|  | **Phenotype IV** | **Non-phenotype IV** |  |
| Age (years) | 74 ± 11 | 64 ± 14 | **0.003** |
| LV EF 35-50% | 50% | 5% | **<0.001** |
|  | **Phenotype V** | **Non-phenotype V** |  |
| MAP (mmHg) | 57 ± 11 | 75 ± 16 | **<0.001** |
| Creatinine (mg/dl) | 6.26 (5.40-8.81) | 1.40 (1.01-2.08) | **<0.001** |

Values are expressed as mean ± SD or count and percentage or median (IQR). EDD, end-diastolic diameter; EF, ejection fraction; LV, left ventricular; MAP, mean arterial pressure; RV FAC, right ventricular fractional area change; S' TDI, s' wave tissue doppler imaging.

As for CS etiology, the majority of patients in Phenotype I, IV and V had ACS-related CS, accounting for 72%, 71%, and 80% of cases, respectively. In contrast, 82% of patients with Phenotype II and 51% with Phenotype III had non-ACS-related CS. No statistically significant difference regarding the etiology was observed between Phenotype I and Phenotype IV (P = 0.842) or Phenotype V (P = 0.640).

Finally, no statistically significant differences were found in the use of MCS across all the Phenotypes (P = 0.332; specifically: P = 0.149 for IABP; P = 0.653 for Impella; P = 0.199 for ECMO; P = 0.767 for ECMO + IABP; P = 0.306 for ECMO + Impella).

### ASSOCIATION OF PHENOTYPES WITH OUTCOME

In-hospital mortality rates progressively increased across the phenotypes, with rates of 25%, 32%, 39%, 41% and 60% for Phenotype I through V, respectively (*Figure S4*). Univariate Cox regression analysis revealed a statistically significant rise in mortality corresponding to higher phenotype classification. Cox regression analysis also revealed that, relative to Phenotype I, patients with Phenotype V and IV were at highest risk of mortality (hazard ratio (HR): 2.78 [95% confidence interval (CI), 1.07-7.19] and HR: 2.80 [95% CI, 1.10-7.14] respectively). At multivariate analysis, Phenotypes predicted mortality also after adjusting for confounding factors (age, sex, ACS-related etiology, MCS utilization; adjusted HR 1.29, [95% CI, 1.07-1.57, P = 0.009) (*Table 3*).

**Table 3.** Multivariate and Bivariate Cox analysis.

| Multivariate Cox analysis adjusting Phenotype classification for confounding factors. |  |  |  |  |
| --- | --- | --- | --- | --- |
| Variables | B | HR | CI 95% | P value |
| Phenotype | 0.26 | 1.29 | 1.07-1.57 | **0.009** |
| Age | 0.06 | 1.06 | 1.04-1.09 | **<0.001** |
| Female sex | 0.08 | 1.09 | 0.60-1.97 | 0.789 |
| ACS-related etiology | 0.67 | 1.94 | 1.08-3.49 | **0.026** |
| MCS | -0.12 | 0.89 | 0.49-1.59 | 0.697 |
| Bivariate Cox analysis including Phenotype and SCAI stage. |  |  |  |  |
| Variables | B | HR | CI 95% | P value |
| Phenotype | 0.216 | 1.24 | 1.01-1.53 | **0.041** |
| SCAI stage | 0.400 | 1.49 | 1.06-2.09 | **0.021** |

ACS, acute coronary syndrome; CI, confidence interval; MCS, mechanical circulatory support; HR, hazard ratio, SCAI, Society for Cardiovascular Angiography and Interventions.

Survival analysis by Kaplan-Meier showed a stepwise increase in mortality for the five clusters (*Figure 4*); the log-rank tests between all survival curves of the different Phenotypes showed a P = 0.109, but a statistically significant difference in cumulative survival between Phenotype I and Phenotype V (Chi-square 4.880, P = 0.027) or Phenotype IV (Chi-square 5.652, P = 0.017) was observed.

**Figure 4.**
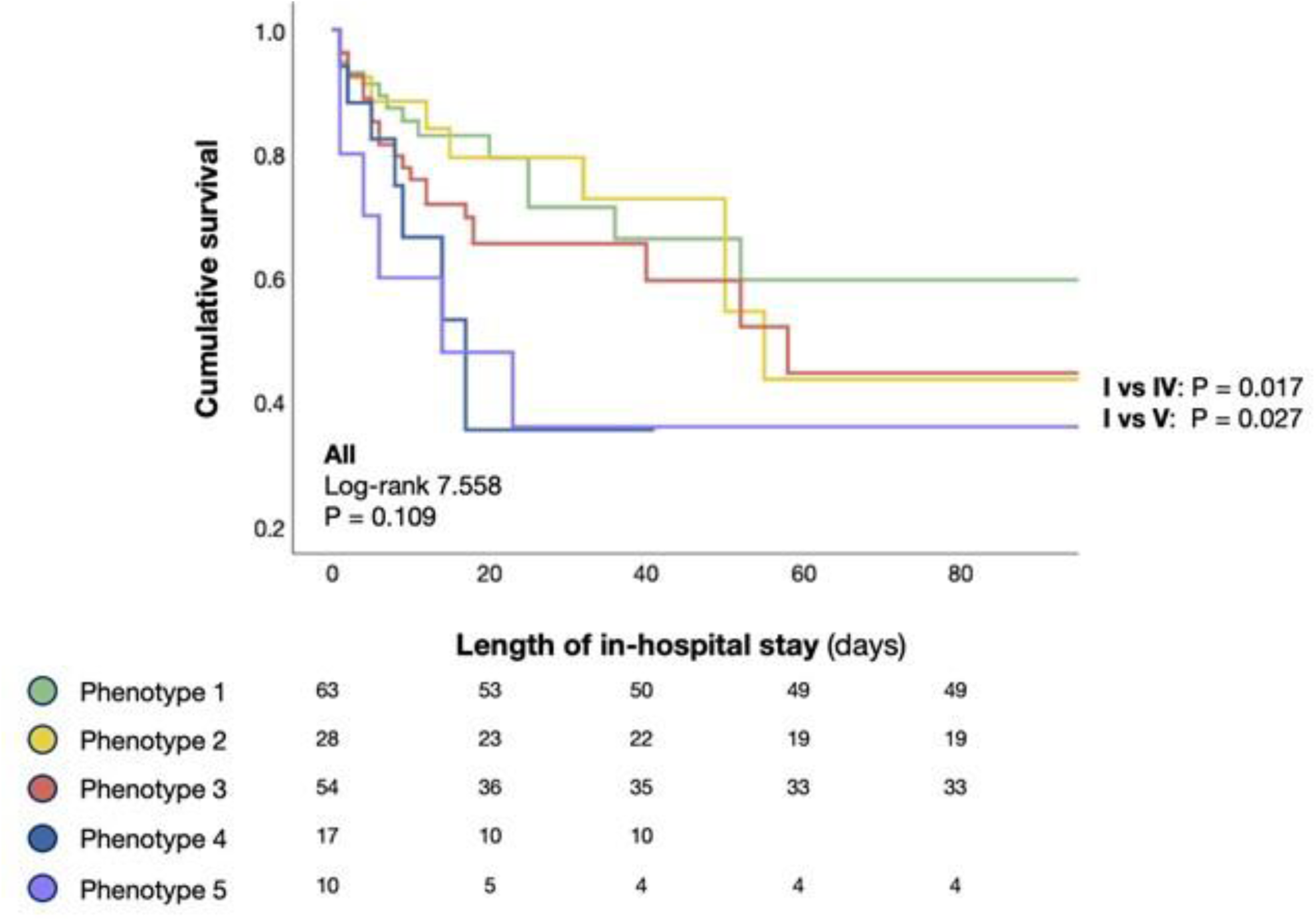
Association of Phenotypes with outcome – Kaplan-Meier survival curves for the 5 cardiogenic shock Phenotypes.

### PHENOTYPES AND SCAI STAGES

We compared the distribution of each Phenotype with regard to their SCAI shock stage at admission (*Figure 5*): within each Phenotypes, the SCAI staging further stratified mortality. Moreover, within each SCAI stage (C-E), the 5 Phenotypes further stratified mortality. Eventually, at bivariate Cox regression analysis, Phenotypes predict mortality independently of SCAI stage (HR 1.24, CI 1.01-1.53, P = 0.041) (*Table 3*).

**Figure 5.**
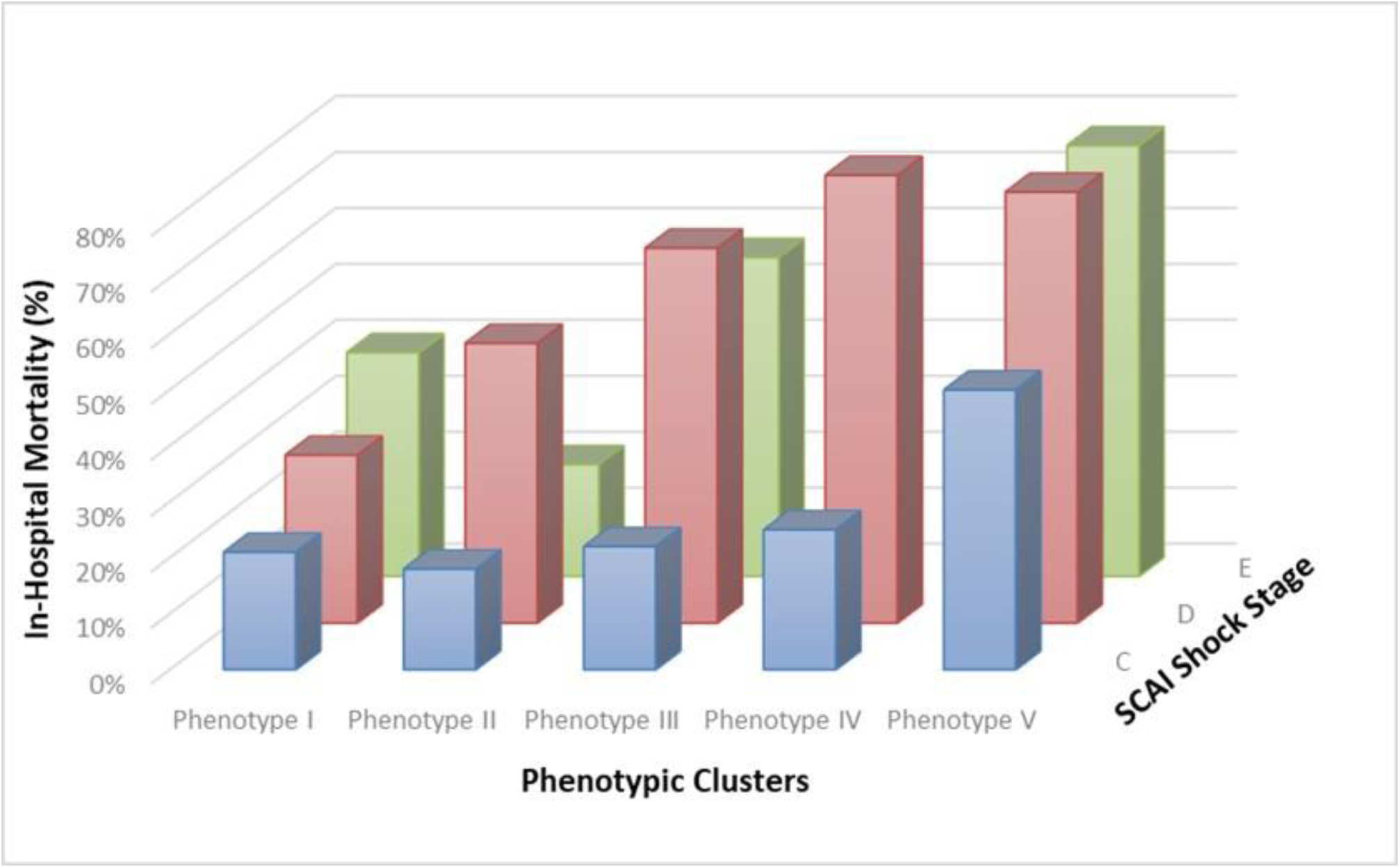
Phenotypes and SCAI stages - In-hospital Mortality according to the SCAI Shock Classification and cardiogenic shock Phenotype. SCAI, Society for Cardiovascular Angiography and Interventions.

## DISCUSSION

To date, this is the first prospective, multicenter study to identify CS sub-phenotypes through a ML algorithm, using not only clinical variables, but also echocardiographic data, beyond etiology.

These Phenotypes differ in hemodynamic and metabolic characteristics, as well as in cardiac structure and function, suggesting the presence of distinct clinical groups that are expected to correlate with in-hospital mortality, providing a foundation for personalized treatment strategies in CS patients (all distinctive characteristics of Phenotypes have significant P values). The widespread availability and rapid execution of echocardiography allow for the seamless integration of this information, making this phenotyping approach easily feasible at bedside. In this direction, our results showed that marked cardiac dysfunction and dilatation were not a major discriminator of worse prognosis across all the Phenotypes, according with limited studies that demonstrated the lack of association between severe LV dysfunction and mortality [24–26].

Considering a total of 172 patients, the population was significantly representative of overt CS (SCAI stage C-E), and 65% patients received at least one mechanical device for cardiac/circulatory support, ranging from intra-aortic balloon pump to extracorporeal membrane oxygenators. Phenotype I, that showed the lowest mortality and predominantly ACS-related etiology, had a higher mean arterial pressure, normal kidney function and a slight increase in LV diameter with a moderate reduction in EF and LV GLS. Phenotype II, also associated with lower mortality, exhibited similar metabolic and hemodynamic features but, interestingly, marked LV dilation and dysfunction and normal right ventricular function, identifying younger patients probably with a history of dilated cardiomyopathy. Phenotype III primarily identified CS patients of intermediate age and in-hospital mortality, with relevant RV dysfunction combined with marked systemic congestion and metabolic and renal impairment. Otherwise, Phenotype IV mainly detected older and ACS-related CS patients associated with the highest mortality, alongside Phenotype V. In addition, Phenotype V was distinguished by a marked state of hypotension associated with a potentially consequent severe renal dysfunction.

Many of the phenotypes’ clinical characteristics align with established clinical scores and classifications, reinforcing the concepts that hemodynamic and metabolic parameters are associated with increased mortality [8, 9, 12, 28–32]. Either Phenotypes IV and V showed relevant metabolic impairment and systemic congestion, but only a mild reduction in LV EF with normal LV dimension and right ventricular function, suggesting that cardiac function differs between Phenotypes and metabolic derangements can occur in the absence of a marked reduction in LV EF. In this sense, it is becoming increasingly necessary to consider circulatory shock – thus including CS – as a syndrome that originates from hemodynamic alterations of various causes but progresses toward a common final pathway of metabolic derangement, ultimately evolving into a cellular and mitochondrial pathology that influences outcome.

Moreover, K. Berg-Hansen et al. demonstrated in a recent study that LV GLS proved as a superior predictor of short-term mortality in patients with CS compared with LV EF (which was not associated with in-hospital mortality), indicating that regular LV GLS assessment could improve risk stratification in this cohort [21]. In our study, clustering was also performed using LV GLS as a variable to evaluate functional cardiac alterations across different Phenotypes. The results revealed that LV GLS impairment occurred alongside a decline in LV EF; however, since the higher sensitivity, LV GLS could give an earlier ability to predict overall prognosis.

In addition, these subgroups transcend the concept of etiology-based phenotyping, which remains difficult due to the inherently heterogeneous population even within ACS-related CS category. Indeed, the correlation between Phenotypes and in-hospital mortality is independent of the CS etiology.

Phenotypes could also provide support to SCAI staging system: in our analysis, within each Phenotypes, the SCAI staging further stratified mortality, even if SCAI stage E in Phenotypes II, III and IV was relatively underrepresented; this may be at least partially explained by the fact that SCAI classification is dynamic and patients may progress to higher stages during hospital stay [26]. Furthermore, within each SCAI stage (C-E), the 5 Phenotypes further stratified mortality, and Phenotypes predicted mortality independently of SCAI stage, even after adjustment for confounding factors. Hence, the Phenotypes seem to be compatible with the SCAI stage at admission and may provide further support for it in more personalized therapy.

Clustering methods have been effectively utilized for phenotyping CS patients in different studies: Soussi S et al. individuated four biomarker-driven CS sub-phenotypes [33], while Zweck et al. employed K-means algorithm to identify three subgroups based on 6 admission laboratory selected variables. The three clusters individuated were labeled as “non-congested,” “cardiorenal,” and “cardiometabolic” subgroups based on their clinical characteristics [18, 19, 34]. Using K-means clustering to sub-phenotype patients with CS could offer some advantages: one of the main strengths of K-means lies in its simplicity and interpretability, making it an accessible tool for exploratory analysis. The algorithm’s computational efficiency is particularly well-suited to datasets of moderate size, such as in this study. Moreover, as an unsupervised method, K-means does not require labeled data, which is particularly advantageous when the goal is to uncover hidden patterns or identify novel phenotypes based on clinical features. Using this method, we found 5 Phenotypes based not only on their clinical variables (as in previous studies), but also on echocardiographic characteristics at presentation which differ from each other, suggesting the individuation of distinct clinical clusters.

### STUDY LIMITATIONS

Major limitations of this study include the limited number of patients and clinical variables, the inability to assess long-term outcomes and the lack of an external validation to other independent CS cohorts, which is important to assess the reproducibility of the analysis. Further larger prospective studies that account for temporal changes in patients’ parameters are needed, to capture the potential transition between Phenotypes during hospitalization. Moreover, the ML method assumes that clusters are spherical and evenly distributed in the feature space, which may not align with the complex, heterogeneous nature of clinical data. Furthermore, the algorithm is sensitive to outliers, which can disproportionately impact the positioning of cluster centroids, and its limited capacity to handle high-dimensional data effectively without preprocessing. Finally, the dependence of speckle-tracking echocardiography on image quality should be considered, reflecting daily practice challenges in cardiac intensive care unit imaging, though sensitivity analysis confirmed the robustness of clustering results even when LV GLS was excluded.

## CONCLUSIONS

We identified 5 CS Phenotypes through a ML approach, combining echocardiographic data with metabolic and hemodynamic variables. These subgroups, characterized by unique mortality risks and features, including varying patterns of cardiac dysfunction and dilatation, offer a framework for tailoring treatment strategies and improving risk stratification beyond SCAI staging in CS patients.

## Data Availability

The corresponding author had full access to all the study data and takes responsibility for its integrity and the data analysis.

## Sources of Funding

no funding related to this work.

## Disclosures

none.

## Supplemental Material

Supplemental Methods

Figure S1-S4

## Nonstandard Abbreviations and Acronyms

ACS: acute coronary syndrome
CS: cardiogenic shock
EDD: end-diastolic diameter
EF: ejection fraction
GLS: global longitudinal strain
LV: left ventricular
MAP: mean arterial pressure
ML: machine learning
SCAI: Society for Cardiovascular Angiography and Interventions
S’ TDI: wave s’ tissue Doppler imaging

## Notes

### Competing Interest Statement

The authors have declared no competing interest.

### Author Declarations

The study was approved by the Tuscany Region Local Ethic Committee for Clinical Trial – section south-west extended area (approval number: 20438).

